# Behavioral Factors Associated with Blood Pressure Control in Hypertensive Adults 35 to 64 Years of Age at Risk of Future Stroke: A cross-sectional survey study

**DOI:** 10.1101/2023.08.21.23294394

**Authors:** Patricia Railsback Masson, Barbara Ellen Mawn, Thabele (Bay) Leslie-Mazwi, Kristen A. Sethares, Alyson Caruso, Jason Rydberg

**Affiliations:** Massachusetts General Hospital Boston, MA; University of Massachusetts Lowell, Solomont School of Nursing, Lowell, MA; Department of Neurology, University of Washington Medicine, Seattle, WA; University of Massachusetts Dartmouth, Dartmouth, MA; Dana Farber Cancer Institute, Boston, MA; University of Massachusetts Lowell, College of Fine Arts, Humanities and Social Sciences, Lowell, MA

## Abstract

**BACKGROUND:** The incidence of stroke has increased for those < 64 years of age over the last decade. Control of hypertension is one of the greatest modifiable risk factors for preventing stroke but remains elusive. In 2017, the American College of Cardiology and American Heart Association national blood pressure guidelines tightened the definition of a normal blood pressure to <120/80 mm Hg, which if achieved would reduce stroke incidence by up to 21%.

**METHODS:** This IRB approved cross-sectional study utilized stratified random sampling to recruit adults, 35 to 64 years of age with an active diagnosis of hypertension, no prior history of stroke or transient ischemic attack (TIA) and under current primary care. Blood pressure was dichotomized into controlled blood pressure (≤120/80 mm Hg) and uncontrolled blood pressure (>120/80 mm Hg). Guided by a modified Health Belief Model, electronic surveys assessing individual medication compliance and beliefs about future stroke risk were distributed. Bivariate and multivariate logistic regression models analyzed the data with α set at 0.05 and observational cohort guidelines were followed.

**RESULTS:** Of the 304 subjects, 83% (n=253) had uncontrolled blood pressure, 78% of women and 89% of men, (77% of total subjects) classified as stage 1 or stage 2 hypertension. Men and women had mean composite compliancy scores below the 50^th^ percentile (21.53 +/-2.89). Women with uncontrolled hypertension were statistically less likely to be compliant (*p*= 0.040), with lower perceived susceptibility (*p= 0.04*) and severity of stroke (*p=0.037*) and less likely to see barriers to initiating exercise (*p=0.04)* but did not view exercise as beneficial in preventing stroke (*p*=0.039).

**CONCLUSION:** Uncontrolled blood pressure remains problematic for both men and women. Women are known to have greater longevity and this study has identified targeted beliefs and behaviors that may more effectively address uncontrolled blood pressure in women at risk for future stroke.

## Introduction

It is estimated that 116 million American adults have hypertension. ^1^ Hypertension is one of the greatest risk factors for stroke and outcomes for stroke prevention measures have worsened in the last five years especially among adults less than 65 years of age. ^1,2,3^ Blood pressure control is estimated to be achieved in only 23% of these 116 million individuals. Reducing blood pressure to <120/80 mm Hg has been associated with a 21% reduction in stroke and an overall 10-year cardiovascular (CVD) risk reduction. ^4, 5^ The American Centers for Disease Control and Prevention and American Heart & Stroke Association’s Million Hearts^©^ 2022 program was launched in 2018 with the goal of improving blood pressure control among hypertensive adults, ages 18 and older, from 16% to 18%. This goal was not met in 2022 and the campaign has been extended for an additional five years. ^1, 2, 3^ Hypertensive adults 35 to 64 years of age are identified in the campaign as a priority population because of rising stroke rates in this population. ^3^

A variety of factors influence blood pressure control. Individuals who are actively engaged in treatment and monitoring of hypertension are more likely to have greater hypertension knowledge and report higher treatment compliance but not necessarily have blood pressure control. ^6, 7, 8, 9, 10, 11^ Research has demonstrated that individual blood pressure control may be influenced by other associated risk factors such as sex, age, race/ethnicity, socioeconomics, diabetes, hyperlipidemia, heart conditions, smoking, diet, exercise and stress, all risk factors commonly associated with stroke. ^3, 4, 12, 13, 14, 15, 16^, However, behavioral factors are poorly understood and explored. This is especially true for younger patients, because the majority of literature to date has focused on adults older than 65 years of age and those who have already experienced a TIA or stroke. The aims of this study were to assess blood pressure control and evaluate the population of hypertensive adults 35 to 64 years of age to explore behavioral factors that influence blood pressure control.

## Methods

This IRB approved, cross-sectional study used patient portals, electronic medical records (EMR) and validated survey tools to obtain blood pressure control status, self-reported treatment compliance, beliefs and behaviors in hypertensive adults 35 to 64 years of age. A northeast healthcare system data base was accessed to identify 1,200 potential subjects and stratified random sampling was employed to maximize equitable sex and age (male and female, 35-44, 45-54 & 55-64) distribution. Study eligibility required: ages 35 to 64, active diagnosis of hypertension, not pregnant, under established primary care and no current or past diagnosis of transient ischemic attack (TIA) or stroke. Three hundred-nine surveys were completed between January to April of 2022, five were excluded due to past stroke or incomplete behavioral survey data resulting in 304 (25.3%) participants.

### Protection of Human Subjects

Study invitational letters were sent via eligible patient portals. Once participants agreed to participate in response to the first letter, a second letter was sent securely to their preferred email. This letter gave additional details about the study and provided an embedded secure electronic link to the healthcare system’s HIPAA compliant secure REDCap tool^©^. This study was voluntary; individuals were informed that they did not have to participate. Survey completion implied consent and only completed surveys (consent) were used to complete the EMR data abstraction. All data were secured electronically through the healthcare system’s electronic platforms.

### Blood Pressure Measurement

Blood pressure control parameters, as established by the American College of Cardiology and the American Heart Association were utilized: Normal (<120/<80 mmHg), Elevated (120-129/<80 mmHg), Stage 1 (130-139/or 80-89 mmHg), Stage 2 (≥140/or >90 mmHg) and Hypertensive Crisis (>180/&/or>120 mmHg). Blood pressure readings were obtained from the most recent routine primary care visit only. For the purpose of this study and evidence supporting lower blood pressure ranges to mitigate 10-year CVD risk and stroke prevention, blood pressure control was dichotomized into controlled blood pressure, normal:<120/80 mm Hg and uncontrolled blood pressure, >120/80 mm Hg (elevated, stage 1, stage 2, and hypertensive crisis). ^1, 4, 5^

### Research Theoretical Model and Survey Tools

The Health Belief Model (HBM) has been effectively used in numerous studies assessing health beliefs and risk factors for stroke and blood pressure control. ^8, 9, 12, 15, 17, 21, 22, 23, 24^ For this study, the HBM was modified to evaluate individual beliefs about severity and susceptibility to a future stroke and behaviors such as exercise as a means to control blood pressure and prevent future stroke.

### Survey Tools

Permission was obtained to use the Hill-Bone Compliance to High Blood Pressure Therapy Scale (HB-HBPS) ^25^ and the Cerebrovascular Attitudes and Beliefs Scale-Revised Scale (CABS-R). ^26^ The HB-HBPS examines three key areas of individual behavior: medication compliance, appointment keeping and sodium intake, allowing for subgroup analysis and a composite score with a reported α of 0.74 to 0.84 and inter-item correlation of 0.18 to 0.28. ^25^ The CABS-R examines individual beliefs of stroke severity and susceptibility and the benefits and barriers of exercise as a means to prevent stroke with a reported α of 0.79 to 0.89 and inter-item correlations of greater than 0.15. ^26^ Additionally, subjects were asked what they believed their chances were of having a stroke in the future.

### Statistical Analysis

Blood pressure control was dichotomized into controlled blood pressure and uncontrolled blood pressure. Predictor variables included: sex, age, race/ethnicity, education, income, marital status, HB-HBPS, CABS-R and the chance of future stroke score. Missing data (income 10, education 2 and marital status 3) were mode imputed. A G*Power analysis with a medium effect and β 0.05 required 251 participants to be enrolled. Analyses were conducted in the R statistical computing environment and included; descriptive statistics, distribution curves, means and odds ratios (OR) obtained from multiple logistic regressions.

## Results

Of the 304 participants, 53% (n=162) were women, 83% were white (n=251), 69% (n=210) were college educated, 50% (n=146) had incomes greater than $140,000/year and 64% (n=193) were married. Eighty-three percent (n=253) of participants (89%, n=126 men and 78%, n=127 women) in this study had uncontrolled hypertension, with 77% (n=194) of these individuals classified as Stage 1 or Stage 2 hypertension. See Table 1.

**Table 1.** Population Demographics & Blood Pressure Control.

|  | **Men**<br>**n=142** |  | **Women**<br>**N=162** |  | **Total**<br>**n=304** |  |
| --- | --- | --- | --- | --- | --- | --- |
|  | n | % | n | % | n | % |
| **Age** |  |  |  |  |  |  |
| 35-44 | 40 | 13.2 | 50 | 16.5 | 90 | 29.6 |
| 45-54 | 46 | 15.1 | 54 | 17.8 | 100 | 32.8 |
| 55-64 | 56 | 18.4 | 58 | 19.1 | 114 | 37.5 |
|  | 142 | 46.7 | 162 | 53.3 | 304 | 100 |
| **Race** |  |  |  |  |  |  |
| Non-white | 20 | 14.1 | 33 | 20.4 | 53 | 17.0 |
| White | 122 | 85.9 | 129 | 79.6 | 251 | 83.0 |
| **Income (n=294)** |  |  |  |  |  |  |
| <\$50,000 | 23 | 16.7 | 28 | 17.9 | 51 | 17.0 |
| \$50,000-\$140,000 | 37 | 26.8 | 60 | 38.4 | 97 | 33.0 |
| >\$140,000 | 78 | 56.5 | 68 | 43.6 | 146 | 50.0 |
| **Education (n=302)** |  |  |  |  |  |  |
| Some college or less | 41 | 28.9 | 51 | 31.9 | 92 | 30.0 |
| Associates or Bachelors | 57 | 40.1 | 61 | 38.1 | 118 | 40.0 |
| Masters or Doctorate | 44 | 30 | 48 | 30.0 | 92 | 30.0 |
| **Marital Status** |  |  |  |  |  |  |
| Married (currently) | 92 | 64.8 | 101 | 63.5 | 193 | 64.0 |
| Not Married | 50 | 35.2 | 58 | 36.5 | 108 | 36 |
| **Blood Pressure Status** |  |  |  |  |  |  |
| **Controlled Blood Pressure** |  |  |  |  |  |  |
| Normal (<120SBP/<80DBP mmHg) | 16 | 11.0 | 35 | 22.0 | 51 | 17.0 |
| **Uncontrolled Blood Pressure** |  |  |  |  |  |  |
| Elevated (120-129SBP/<80DBP mm Hg) | 31 | 22.01 | 23 | 14.0 | 54 | 18.0 |
| Stage 1 (130-139SBP/80-89DBP mm Hg) | 47 | 33.0 | 56 | 35.0 | 103 | 34.0 |
| Stage 2 (≥140SBP &/or >90DBP mm Hg) | 48 | 34.0 | 43 | 27.0 | 91 | 30.0 |
| Crisis (>180SBP &/or >120DBP mm Hg) | 0 | 0.0 | 5 | 2.0 | 5 | 1.0 |

### Individual Beliefs of Future Stroke Risk

Overall, individuals with uncontrolled blood pressure were more likely to believe they were at risk for a future stroke, however, these differences are estimated with considerable uncertainty between groups (OR 1.5; 95% CI: 0.86, 2.69). Women with uncontrolled blood pressure were less likely to believe they would have a stroke in the future OR 0.49 (95% CI: 0.25, 0.93; *p=0.034*) than men with uncontrolled blood pressure and women with controlled blood pressure.

### Hill Bone-High Blood Pressure Score (HB-HBPS)

The HB-HBPS mean composite score ranges from 14 to 56 and both controlled (21.529 +/-2.89) and uncontrolled (21.877 +/-3.37) blood pressure groups had mean composite scores below the 50^th^ percentile. While there was little difference between the blood pressure control groups, women with uncontrolled blood pressure were less likely to have medication compliance (OR 0.50; 95% CI: 0.25, 0.95; *p*=0.039), keep appointments (OR 0.40; 95% CI: 0.25, 0.94; *p*= 0.03) or manage/reduce sodium intake (OR 0.49; 95% CI: 25, 0.94; *p*=0.036) than men with controlled and uncontrolled blood pressure.

### Cerebrovascular Attitudes & Beliefs Scale-Revised (CABS-R)

The CABS-R like the Health Belief Model negates composite scoring as it could reduce or dilute the effect of each subscale. *Perceived Severity of Stroke (range 3-15):* Both blood pressure control groups scored above the 75^th^ percentile for perceived severity of stroke (13.24 +/-1.93, 13.25 +/-1.80). Participants with uncontrolled blood pressure who graduated from college OR 0.44 (95% CI: 0.11, 1.48), had incomes < $140,000/annually OR 0.74 (95% CI: 0.25,2.00) and were married OR 0.82 (95% CI: 0.39, 1.65) were less likely to perceive stroke as severe, however, similarity between groups could not be ruled out. Women with uncontrolled blood pressure were less likely to perceive stroke as severe, OR 0.5 (95% CI: 0.25, 0.95; *p=* 0.037) when comparing to men with controlled and uncontrolled hypertension. *Perceived Susceptibility to Stroke (range: 4-16):* Participants with controlled blood pressure had slightly higher mean scores (9.705 +/-2.39) for perceiving their susceptibility to stroke than those with uncontrolled blood pressure (9.40 +/-2.33), however, both groups were below the 50^th^ percentile.

Women with uncontrolled blood pressure were less likely to perceive being susceptible to having a stroke, OR, 0.5 (95% CI: 0.25, 0.96; *p*=0.04). *Perceived Benefits of Exercise (range: 8-40):* Participants from both groups scored above the 50^th^ percentile for believing exercise is a way to prevent stroke (controlled blood pressure 28.69 +/-4.03, uncontrolled blood pressure 28.92 +/-3.95). However, women with uncontrolled blood pressure, OR 0.5 (95% CI: 0.25, 0.95; *p*=0.039), individuals with income <$140,000/annually OR 0.74 (95% CI: 0.25, 2.00), were married OR 0.82 (95% CI: 0.39, 1.65) and college educated OR 0.42 (95% CI: 0.12, 1.21) were less likely to perceive the benefits of exercise as a means to prevent stroke. *Perceived Barriers to Exercise (range: 8-40):* Participants from both groups perceived barriers to exercise with mean scores in the 50^th^ to 75^th^ percentile (controlled blood pressure 29.76 +/-3.25, uncontrolled blood pressure 28.90 +/-3.81). Women with uncontrolled blood pressure were less likely to perceive barriers to exercise, OR 0.51 (95% CI: 0.26, 9.84e-01; *p*=0.049) and men with controlled and uncontrolled blood pressure were more likely to perceive barriers to exercise if they were white OR 1.38 (95% CI: 0.55, 3.29e+00) or had incomes >$140,000/annually OR 1.06 (95% CI: 0.31, 3.34e+00). See Table 2.

**Table 2.** Population Stroke Risk Belief HB-HBPS & CABS-R Scores.

|  | Uncontrolled Blood Pressure |  |  |  |  |
| --- | --- | --- | --- | --- | --- |
|  | Mean | OR<br>(odds ratio) | SE | 95% CI<br>(Confidence<br>Interval) | p-value |
| **Future Stroke Risk Belief - sample** |  | 1.5 | 0.29 | 0.86, 2.69 | 0.17 |
| Women |  | 0.49 | 0.34 | 0.25, 0.93 | 0.034 |
| **Hill Bone – High Blood Pressure Scores** |  |  |  |  |  |
| Composite (14-56) | 21.88 +/- 3.37 | 1.03 | 0.05 | 0.94, 1.15 | 0.54 |
| Composite – Women |  | 0.5 | 0.34 | 0.25, 0.95 | 0.04 |
| Appointment Keeping (2-8) | 3.14 +/- 1.05 | 1.12 | 0.16 | 0.82, 1.54 | 0.49 |
| Appointment Keeping – Women with uncontrolled BP |  | 0.49 | 0.34 | 0.25, 0.94 | 0.03 |
| Medication Taking (9-36) | 13.34 +/- 2.64 | 1.05 | 0.07 | 0.93, 1.24 | 0.45 |
| Medication Taking – Women with uncontrolled BP |  | 0.5 | 0.34 | 0.25, 0.95 | 0.039 |
| Sodium Intake (3-12) | 5.4 +/- 1.26 | 0.94 | 0.12 | 0.74, 1.21 | 0.61 |
| Sodium Intake – Women with uncontrolled BP |  | 0.49 | 0.34 | 0.25, 0.94 | 0.036 |
| **Cerebrovascular Attitudes & Beliefs Scale - Revised** |  |  |  |  |  |
| Severity of Stroke (3-15) | 13.25 +/- 1.80 | 0.99 | 0.09 | 0.82, 1.18 | 0.95 |
| Severity of Stroke – Women with uncontrolled BP |  | 0.5 | 0.34 | 0.25, 0.95 | 0.037 |
| Susceptibility to Stroke (4-16) | 9.40 +/- 2.33 | 0.94 | 0.07 | 0.81, 1.08 | 0.37 |
| Susceptibility to Stroke – Women with uncontrolled BP |  | 0.5 | 0.34 | 0.25, 0.96 | 0.04 |
| Benefits of Exercise to Prevent Stroke (8-40) | 28.92 +/- 3.95 | 1.0 | 0.04 | 0.92, 1.09 | 0.99 |
| Benefits of Exercise – Women with uncontrolled BP |  | 0.5 | 0.34 | 0.25, 0.95 | 0.039 |
| Barriers to Exercise to Prevent Stroke (8-40) | 28.80 +/- 3.81 | 0.92 | 0.05 | 0.84, 1.00e+00 | 0.07 |
| Barriers to Exercise – Women with uncontrolled BP |  | 0.51 | 0.34 | 0.26, 9.84e-01 | 0.049 |

## Discussion

Blood pressure control is a key factor in preventing future stroke in any population. Individual beliefs and clinician disagreements on how to achieve optimal control potentially contribute to many individuals with hypertension not receiving recommended blood pressure treatment, including escalation of therapies ^17, 18, 19^ This study focusing on a priority age group for blood pressure control, demonstrated that blood pressure control remains a challenge with 17% (n=51) of the participants classified as having a normal blood pressure (≤120/80 mm Hg) compared to the national estimate of 26.1%. ^27^ These findings are especially concerning because they occur in populations previously considered to be at lower risk for uncontrolled blood pressure, white, educated, higher income earners under active primary care in a comprehensive coordinated healthcare system.

### Population Compliance

Our current model of care is dependent on healthcare providers to identify and share with patients the clinical risk factors and courses of actions to be taken to reduce or manage the risks of hypertension. Individuals may be given advice, medications and follow-up care visits that could be weeks, months or a year later. ^5, 17, 19, 28^ Primary care clinicians have the burden of identifying hypertension and establishing treatments, but are often faced with heavy workloads, lack of trust in blood pressure measurements in and out of clinic settings and reluctance to treat or escalate hypertension treatment in younger populations. ^17, 18, 19, 20^ Once risk factors and treatment plans are established, it then becomes dependent on the individual to actively comply. All patients in this cohort already carried a diagnosis of hypertension, and were in established primary care, overcoming one of the identified barriers in blood pressure control of healthcare provision. Given the sociodemographic profile of this cohort (predominantly white, well-educated with high incomes) compliance among controlled and uncontrolled participants was lower than had been expected. Closer examination of the subscale scores revealed that, medication compliance, sodium intake and appointment keeping scores all fell below the 50^th^ percentile and this was even more relevant for women.

### Population Beliefs

The modified HBM for stroke risk developed for this study posits that individuals with hypertension would believe they were susceptible to a future stroke, that stroke is severe and that the benefits of exercise, medication, appointment keeping and sodium intake would outweigh the barriers to initiating exercise and overall treatment compliance . However, both controlled blood pressure and uncontrolled blood pressure participants in this study, while scoring higher or above the mean in belief that stroke is severe (3-15), 13.24+/-1.93 and 13.25+/-1.80, susceptibility to stroke and benefits of exercise beliefs, were found to be in the lower quartiles and barriers to exercise in the upper quartile. These belief scores may be more reflective of a younger population sample, 35 to 64 years of age that are bearing the burdens of caring for families, changing demands of active employment, and unaware of the cumulative effects of long-term uncontrolled hypertension.

### Limitations

Our study is subject to the typical limitations of a survey-based study. Our response rate of over 25% is typical or slightly better than average survey-based studies. The survey approach does allow assessment of individual behavioral motivators. The population surveyed was from a metropolitan area in the Northeast USA and is predominantly white, educated, wealthy and able to access healthcare and were dichotomized into to only two genders. We are concerned that data from populations that are already defined as greater risk for uncontrolled hypertension (socioeconomically, geographically, racially, ethnically, and genders) will demonstrate even more concerning levels of blood pressure control. This remains to be explored in future studies. We elected to utilize the most recent clinic blood pressure recording and therefore do not have temporal or home blood pressure data. This was a pragmatic decision informed by the understanding that “white coat hypertension” is predictive of cardiovascular and stroke risk and associated with chronic hypertension in medical settings also. ^19, 20^

## Conclusion

Blood pressure control continues to be a challenge in younger adult populations 35 to 64 years of age. Despite educational efforts, we are unlikely to reach the revised million hearts^©^ 2027 national goals. Further investigation is needed into patient risk profiles and primary care acknowledgement, documentation and treatment planning. This cohort had moderate to low compliance with medication adherence, appointment keeping and monitoring or reducing sodium intake. Participants did generally believe in the benefits of exercise as a way to prevent stroke but, the majority identified barriers that overshadowed benefits. Research has indicated that racial and ethnic minority populations and those with low income are more likely to have uncontrolled blood pressure. However, this study demonstrated poor control amongst a predominantly white, educated, high income sample of adults, ages 35-64, with regular access to primary care. Focused prevention strategies for women under the age of 65 are recommended in light of the findings related to their perceived lower risk of stroke in the future. Closer assessment of hypertensive adults understanding of uncontrolled hypertension and future stroke risk is needed. Specifically, directing these individuals to the American Heart and Stroke Association’s hypertension and stroke prevention educational websites may impact the individuals knowledge, beliefs and behaviors. Additional research among younger adults with hypertension is needed to develop tailored interventions to reduce the risk of future stroke.

## Data Availability

deidentified data is available for review upon request

## Acknowledgements

Karen Sullivan, Ph.D., Professor Clinical Neuropsychology, Queensland, University, Australia, author of the CABS-R and the John Hopkins School of Nursing HB-HBPS use permissions.

## Contribution & Conflicts of Interest

All authors of this manuscript have contributed substantially to this work and have no conflicts of interest to report.

